# Determinants of facility-based childbirth among adolescents and young women in Guinea: a secondary analysis of the 2018 Demographic and Health Survey

**DOI:** 10.1101/2022.04.06.22273508

**Authors:** Fassou Mathias Grovogui, Lenka Benova, Hawa Manet, Sidikiba Sidibe, Nafissatou Dioubate, Bienvenu Salim Camara, Abdoul Habib Beavogui, Alexandre Delamou

## Abstract

**Introduction:** Maternal mortality remains very high in Sub-Saharan African countries and the risk is higher among adolescent girls. Maternal mortality occurs in these settings mainly around the time of childbirth and the first 24 hours after birth. Therefore, skilled attendance in an enabling environment is essential to reduce the occurrence of adverse outcomes for both women and their children. This study aims to analyze the determinants of facility childbirth among adolescents and young women in Guinea.

**Methods:** We used the Guinea Demographic and Health Survey (DHS) conducted in 2018. All females who were adolescents (15 -19) or young women (20-24 years) at the time of their most recent live birth in the five years before the survey were included. We examined the use of health facilities for childbirth and its determinants using multivariable logistic regression, built through the Andersen health-seeking model.

**Results:** Overall, 58% of adolescents and 57% of young women gave birth in a health facility. Young women were more likely to have used private sector facilities compared to adolescents (p<0.001). Factors significantly associated with a facility birth in multivariable regression included: secondary or higher educational level (aOR=1.81; 95%CI:1.20-2.64) compared to no formal education; receipt of 1-3 antenatal visits (aOR=8.93; 95%CI: 5.10-15.55) and 4+ visits (aOR=15.1; 95%CI: 8.50-26.84) compared to none; living in urban (aOR=2.13; 95%CI: 1.40-3.37) compared to rural areas. Women from poorest households were least likely to give birth in health facilities. There was substantial variation in the likelihood of birth in a health facility by region, with highest odds in NZérékoré and lowest in Labé.

**Conclusion:** The percentage of births in health facilities among adolescents and young women in Guinea increased since 2012 but remains suboptimal. Socio-economic characteristics, region of residence and antenatal care use were the main determinants of its use. Efforts to improve maternal health among this group should target care discontinuation between antenatal care and childbirth (primarily by removing financial barriers) and increasing the demand for facility-based childbirth services in communities, while paying attention to the quality and respectful nature of healthcare services provided there.

## INTRODUCTION

Reducing maternal mortality to achieve Sustainable Development Goal 3 (SDG 3) is a major public health challenge. Despite efforts over the past 25 years, maternal mortality remains disproportionately high in low-and middle-income countries where the need to address the unmet need for family planning is most significant, particularly among adolescents [1]. The World Health Organization (WHO) estimated that 12 million adolescents aged 15-19 years give birth each year globally. The problem is most prevalent in sub-Saharan Africa (SSA), with variations between countries [2]. The recent Demographic and Health Survey (DHS 2018) in Guinea reported a fertility rate of 120 live births per 1,000 adolescents aged 15-19 [3]. This rate is higher than the West African average of 115 per 1,000 [4]. Giving birth during adolescence carries a higher risk of adverse outcomes for both the girl and the baby due in part to the mother’s biological and physiological immaturity [5]. An important strategy to reduce these risks is to improve skilled birth attendance in an enabling environment [6]. However, despite this particular need, sub-Saharan adolescents are less likely to give birth in a health facility than older women [7,8].

The prevalence of facility-based childbirth among sub-Saharan adolescents varies across countries. Overall, an estimated 65% of adolescents give birth in a health facility [9]. This ranges from 23% in Chad [10] to 89% in Gabon [11]. In Guinea, 56% of adolescents used health facilities for the most recent live birth during the DHS 2018 recall period [3]. Factors associated with adolescents’ use of facilities for childbirth are not clear across countries. In a study analyzing DHS datasets from 29 SSA countries, Doctor et al. (2018) found that the likelihood of facility-based delivery increases with maternal age [7]. However, Adde et al. (2020), in a most recent analysis including 28 SSA countries found no difference between adolescents and older women in facility-based childbirth [9]. These two studies using a sample of women aged 15-49 years found that urban residence and higher wealth quintile were associated with a higher likelihood of facility-based childbirth in SSA countries [7,9]. In Bangladesh, Shahabuddin et al. (2016) found that the frequency of facility-based childbirth varied across regions [12]. Some studies among women of childbearing age in SSA also found that a secondary or higher educational level was associated with a higher likelihood of facility-based childbirth [9,13]. In addition, Doctor et al. found that women with at least primary education were more likely to deliver in a health facility than woman with no formal education [7]. In Nigeria, Dahiru et al. (2015) found that women whose husbands had at least primary education were more likely to deliver in a health facility [14]. Further, Adde et al. (2020) and Doctor et al. (2018) found that the odds of facility-based childbirth were higher among women who had completed at least one antenatal visit [7,9]. These findings had also been reported by Sanni et al. in Ethiopia in 2018 [13]. In addition, frequent listening to the radio or watching television were also associated with the use of health facilities for childbirth by women of childbearing age [9].

In a systematic review including 27 studies, Mekonnen et al. (2015) found that distance to the health facility and community factors such as the proportion of educated women in the community and the rate of ANC use in the community were associated with the use of maternal health services among adolescent in SSA [15]. In Niger, Rai et al. (2013) found that the education of both spouses, belonging to a social group such as the Deermal/Songhai, and having some decision-making autonomy increased women’s chances of having safe deliveries [16].

Guinea did not achieve the Millennium Development Goal 4 and is unlikely to achieve the Sustainable Development Goal 3.1 by 2030 (SDG 3.1). The government of Guinea is pursuing an agenda to achieve a maternal mortality ratio (MMR) reduction from 576 per 100,000 live births in 2017 [1] to 343 per 100,000 live births by 2024 [17]. The country has been implementing a policy of free maternal and neonatal emergency health care in public health facilities since 2011 [18]. Adolescents are a priority target for the reduction of maternal mortality and morbidity in the country. However, there are no specific efforts or strategies targeting adolescents [17]. Factors that influence the choice of location for childbirth among adolescents in the context of free maternal health services in Guinea are not well understood. The objective of this study is to estimate the levels and determinants of facility-based childbirth among adolescents and young women aged 20-24 years in Guinea. The results of this study can support the development of targeted strategies to increase the use of maternal health services among this population.

## MATERIALS METHODS

### Data and population

The data used for this study come from the 2018 Demographic and Health Survey (DHS), the fifth to be conducted in Guinea since 1999. The DHS is a cross-sectional nationally representative household survey which used multilevel cluster sampling. Its purpose was to provide comparable data across time and countries on various indicators, including fertility, adult and child mortality, and use of maternal healthcare. The DHS generally uses standard model questionnaires with the possibility of adaptation and addition of optional modules according to country needs. For this study, we included all female respondents residing in sampled household, with at least live birth in the survey’s five-year recall period, if they were 15-19 years (adolescents) or 20-24 years old (young women) at the time of their most recent live birth. Analyses of childbirth location were restricted to the circumstances of the most recent live birth.

### Outcome variable

We used the place of birth as reported by women as the outcome variable. It was categorized into ‘home birth’ (respondent’s home, other home, and other location) and facility-based birth (government hospital; government health center; government health post; private hospital or clinic, and other private medical facility). For a more detailed analysis of health facility type we considered births in the public versus the private sector. Within the public sector, we focused separately on government hospitals which provide the highest level of emergency obstetric and newborn care and serve as referral centers.

### Independent variables

We used Anderson’s Behavioral Model of Health Services utilization [19] (**Fig 1**) to identify relevant and available variables. We included woman’s age at index birth (15-19; 20-24), woman’s highest educational level at survey (no formal education, any primary, and any secondary or higher). The usual age of primary school completion in Guinea is 12 years [20]. We also included woman’s marital status at survey (married/cohabiting or not), women’s religion (Muslim, Christian, other), and number of ANC visits during the pregnancy preceding the index birth (none, 1-3, 4 or more). Women with ‘don’t know’ responses on number of ANC visits were included in the 1-3 category because we assumed that these women had some ANC but could not recall the exact number of visits.

**Figure 1:**
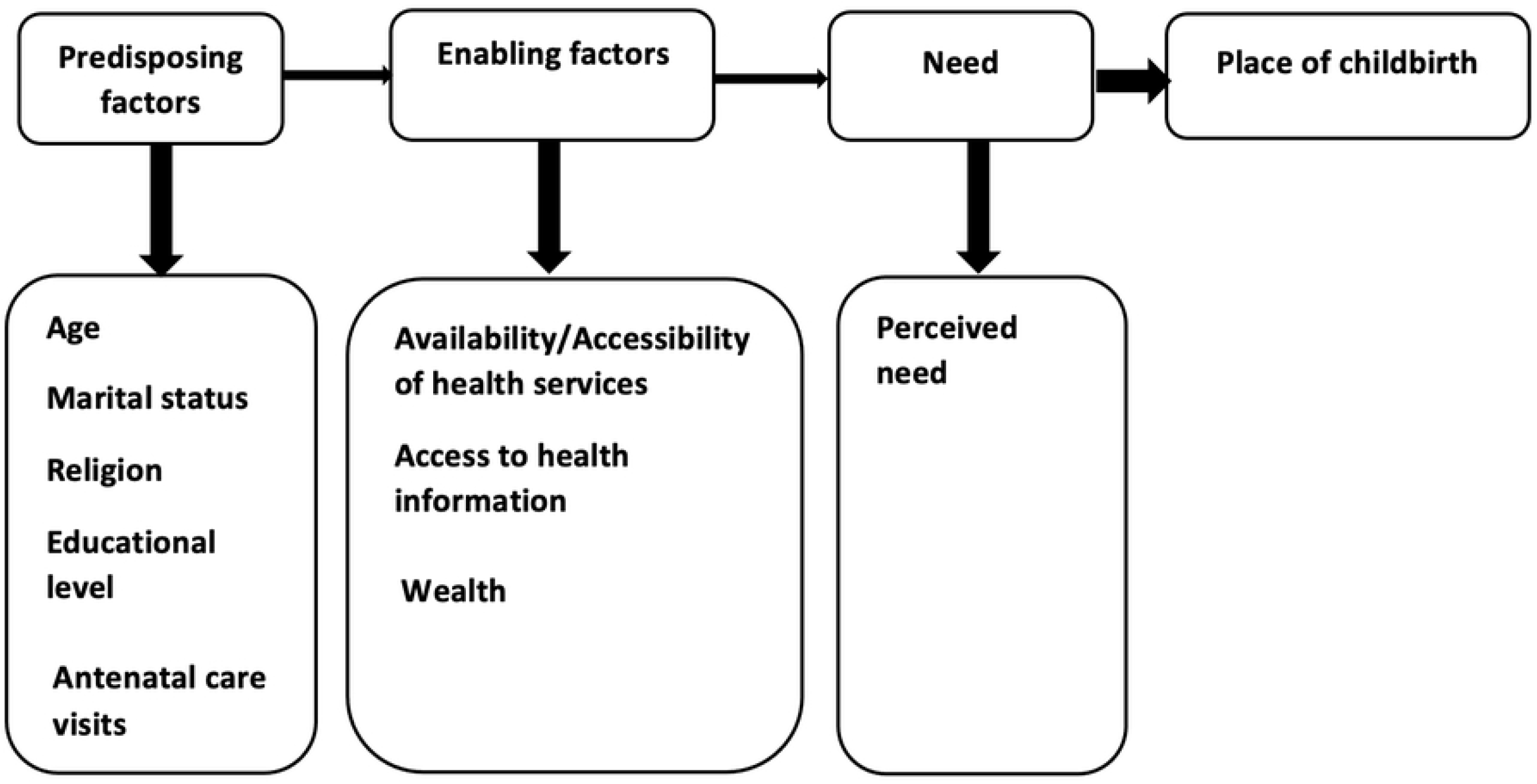
Theoretical framework adapted from Andersen’s Behavioral Model of Health Care Utilization.

Place of residence as specified on the DHS sampling strategy (rural, urban), region of residence (Conakry, Boké, Kindia, Mamou, Labé, Faranah, Kankan, and NZérékoré) and women’s perception of distance to the health facility (big problem or not) were used as proxies for health service availability. Exposure to media (listens to radio or not) was used as a proxy access to health information. We used household wealth quintile (poorest, poorer, middle, richer, and richer) to capture financial access to care. Perceived need factors were mother’s parity (no previous birth, 1 to 2 previous births, 3 or more births) and wantedness of the index pregnancy (wanted, mistimed/unwanted).

### Statistical analysis

The data were processed and analyzed using Stata 16.1 software (StataCorp, College Station, TX USA). Two levels of statistical analysis were applied. The first level consisted of describing the characteristics of the included sample. We obtained absolute numbers and percentages of the characteristics under study for adolescents, young women, and the whole sample. We also compared the prevalence of facility-based childbirth by type of health facility between adolescents and young women using Pearson’s Chi-squared test.

We then analyzed the determinants of facility-based childbirth among the combined sample of adolescents and young women. First, we conducted a bivariate analysis to assess the relationship of each independent variable with adolescents’ or young women’s use of health facilities for their most recent birth. We initially ran one multivariable regression model for each age group, but we found no difference between the characteristics associated with the outcome of interest between the two age groups. Therefore, we constructed a single multivariable model with both age groups. Before fitting the final model, we assessed the level of collinearity between the independent variables. The objective was to determine the relevance of the variables selected for inclusion in the analyses. As a general rule, a mean-variance inflation factor (VIF) score below five is tolerated. In contrast, a mean score greater than or equal to five suggests that the regression coefficients could be misestimated. The VIF reported by the test remained in the tolerable limit.

The Likelihood ratio test (LRtest) was also performed for each variable with more than two categories to assess how its inclusion impacted the over model. Regressions were performed at the 5% threshold, and the results were reported with a 95% confidence interval (95% CI). Differences were considered statistically significant at p<0.05. The Hosmer Lemeshow test was used to test the goodness of fit of the multivariable model. All descriptive and analytical estimated were produced by adjusting for the survey sampling using the svy option, including adjustment for clustering, stratification and unequal probability of selection and non-response (individual sample weights).

### Ethics considerations

A formal request for analysis of all data was made to MEASURE DHS through the online platform, and permission was granted. The original data was collected with ethical approval from the National Ethics Committee for Health Research of Guinea and ICF’s International Review Board.

## RESULTS

### Characteristics of the samples

The sociodemographic characteristics of adolescents and young women included in this study are presented in **Table 1**. We included 934 females who were 15-19 at the time of their most recent live birth in the five years before the survey. Their mean age at the most recent birth was 17.9 years (SD=1.3). Nearly 84% of these adolescents were married/cohabiting, 63% had no formal education, and for 65% this was their first birth. Overall, almost 89% of included adolescents reported at least one ANC visit during their most recent pregnancy, but fewer than two-fifths (36%) received 4 or more ANC visits. The majority of adolescents were Muslim (86%) and lived in rural areas (70%). However, more than half did not perceive the distance to health facility as a big problem (52%).

**Table 1.** Demographic and socioeconomic characteristics of adolescents and young women who had at least one live birth in the five years preceding the DHS 2018 (**N=2**,**154**).

| Characteristics | Adolescents (15-19 years) |  | Young women (20-24 years) |  | Adolescents and young women |  |
| --- | --- | --- | --- | --- | --- | --- |
|  | n=934 | % | n=1,220 | % | n=2,154 | % |
| **Mean age at index birth (years)** | 17.9 (SD=1.3) |  | 22.7 (SD=1.5) |  | 20.6 (SD=1.5) |  |
| **Educational level at survey** |  |  |  |  |  |  |
| No formal education | 578 | 62.7 | 842 | 69.6 | 1420 | 66.6 |
| Primary | 193 | 20.6 | 165 | 12.1 | 358 | 15.8 |
| Secondary or Higher | 163 | 16.7 | 213 | 18.3 | 376 | 17.6 |
| **Marital status at survey** |  |  |  |  |  |  |
| In union/cohabiting | 783 | 83.6 | 1125 | 92.0 | 1908 | 88.4 |
| Not in union | 151 | 16.4 | 95 | 8.0 | 246 | 11.6 |
| **Parity at index birth** |  |  |  |  |  |  |
| No previous birth | 605 | 65.2 | 261 | 21.6 | 866 | 40.5 |
| 1 to 2 previous births | 324 | 34.3 | 760 | 62.5 | 1084 | 50.2 |
| 3 or more previous births | 5 | 0.5 | 199 | 16.0 | 204 | 9.3 |
| **Exposure to media (radio)** |  |  |  |  |  |  |
| Not exposed to media | 362 | 37.5 | 505 | 41.1 | 867 | 39.5 |
| Exposed to media | 572 | 62.5 | 715 | 58.9 | 1287 | 60.5 |
| **Antenatal care visits (ANC)** |  |  |  |  |  |  |
| No ANC | 114 | 11.4 | 152 | 12.1 | 266 | 11.8 |
| 1-3 ANC | 483 | 52.5 | 610 | 49.9 | 1093 | 51.1 |
| 4 plus ANC | 337 | 36.0 | 458 | 38.0 | 795 | 37.1 |
| **Wantedness of index pregnancy** |  |  |  |  |  |  |
| Unwanted or mistimed | 190 | 20.6 | 170 | 14.0 | 360 | 16.9 |
| Wanted | 744 | 79.4 | 1050 | 86.0 | 1794 | 83.1 |
| **Perception of distance to facility** |  |  |  |  |  |  |
| Big problem | 448 | 47.8 | 572 | 44.5 | 1020 | 45.9 |
| not a big problem | 486 | 52.2 | 648 | 55.5 | 1134 | 54.1 |
| **Religion** |  |  |  |  |  |  |
| Muslim | 831 | 86.2 | 1091 | 87.1 | 1922 | 86.7 |
| Christian | 93 | 11.9 | 113 | 11.3 | 206 | 11.5 |
| Other | 10 | 1.9 | 16 | 1.6 | 26 | 1.80.1 |
| **Region** |  |  |  |  |  |  |
| Boké | 136 | 10.4 | 180 | 10.8 | 316 | 10.6 |
| Conakry | 80 | 10.8 | 138 | 14.7 | 218 | 13.0 |
| Faranah | 120 | 10.4 | 150 | 9.1 | 270 | 9.7 |
| Kankan | 160 | 19.7 | 195 | 18.7 | 355 | 19.2 |
| Kindia | 134 | 16.4 | 155 | 14.6 | 289 | 15.4 |
| Labé | 106 | 10.6 | 152 | 11.1 | 258 | 10.9 |
| Mamou | 86 | 6.6 | 96 | 5.9 | 182 | 6.2 |
| NZérékoré | 112 | 15.0 | 154 | 15.0 | 266 | 15.0 |
| **Type of place of residence** |  |  |  |  |  |  |
| Rural | 641 | 69.7 | 813 | 66.4 | 1,454 | 67.9 |
| Urban | 293 | 30.3 | 407 | 33.6 | 700 | 32.1 |
| **Household wealth quintile** |  |  |  |  |  |  |
| Poorest | 202 | 21.5 | 273 | 21.4 | 475 | 21.4 |
| Poorer | 198 | 22.8 | 257 | 21.6 | 455 | 22.1 |
| Middle | 173 | 18.7 | 213 | 17.7 | 386 | 18.2 |
| Richer | 225 | 22.7 | 265 | 21.2 | 490 | 21.9 |
| Richest | 136 | 14.3 | 212 | 18.1 | 348 | 16.5 |

Overall, 1220 young women aged 20-24 at the time of their most recent live birth in the five years before the 2018 DHS were included in the analysis. Their mean age at the most recent birth was 22.7 years (SD=1.5), 92% were married/cohabiting, 63% had two or three previous births, and 70% reported no formal education. Nearly 88% of young women received at least one ANC for their most recent pregnancy, but fewer than two-fifths (38%) completed four or more ANC visits.

### Proportions of facility-based delivery among adolescents and young women

Overall, about 58% adolescents and young women in Guinea gave birth to their most recent baby in a health facility. This proportion was similar in adolescents (58%) and young women (57%). No difference was noted between the two age groups in the use of government hospitals for childbirth (p=0.332). However, young women used private sector facilities more (7.8%) compared to adolescents (3.5%, p<0.001) as shown in **Table 2**.

**Table 2.** Analysis of the frequency of facility-based childbirth by level and health sector among adolescents and young women for the most recent live birth in the five years prior to the 2018 DHS (N=2154).

| Place of childbirth | Adolescents and young women |  | Adolescents (15-19 years) |  | Young women (20-24 years) |  | pvalue |
| --- | --- | --- | --- | --- | --- | --- | --- |
|  | n=2154 | % | N=934 | % | n=1220 | % |  |
| Any health facility | 1209 | 57.6 | 536 | 58.4 | 673 | 57.0 | 0.521 |
| Government hospital | 280 | 12.0 | 129 | 12.8 | 151 | 11.3 | 0.332 |
| Public sector | 1086 | 51.8 | 501 | 55.1 | 585 | 49.3 | 0.012 |
| Private sector | 128 | 6.0 | 37 | 3.5 | 91 | 7.8 | 0.000 |

\*Births in the public sector and private sector together constitute all facility-based births. Government hospital births is a subset of public facility births.

### Determinants of facility-based childbirth among adolescents and young women

**Table 3** shows the percentage of births occurring in health facilities stratified by the independent variables.

**Table 3.** Proportion, bivariate and multivariable analysis of determinants of facility-based childbirth among Guinean adolescents and young women during the five years before the DHS survey in 2018 (n=2,154)

|  | Percentage of facility-based delivery |  |  | Bivariate analysis |  |  |  | Multivariable analysis |  |  |  |  |
| --- | --- | --- | --- | --- | --- | --- | --- | --- | --- | --- | --- | --- |
| Characteristics | % | 95%CI |  | OR | 95%CI |  | Wald test | aOR | 95%CI |  | Wald test | LR test<br>*pvalue |
| Age category |  |  |  |  |  |  |  |  |  |  |  |  |
| 15-19 | 58.4 | 54.4 | 62.3 | 1.06 | 0.89 | 1.27 | **0.521** | 1.05 | 0.80 | 1.37 | **0.746** |  |
| 20-24 | 57.0 | 52.7 | 61.2 | Ref. |  |  |  | Ref. |  |  |  |  |
| Educational level at survey |  |  |  |  |  |  |  |  |  |  |  |  |
| No formal education | 49.7 | 45.3 | 54.1 | Ref. |  |  |  | Ref. |  |  |  |  |
| Primary | 61.8 | 55.5 | 67.8 | 1.64 | 1.21 | 2.22 | **0.002** | 1.09 | 0.80 | 1.58 | **0.640** | **0.004** |
| Secondary or higher | 83.6 | 79.2 | 87.2 | 5.15 | 3.70 | 7.18 | **<0.001** | 1.81 | 1.20 | 2.64 | **0.002** |  |
| Marital status at survey |  |  |  |  |  |  |  |  |  |  |  |  |
| In union/cohabiting | 73.8 | 67.0 | 79.6 | Ref. |  |  |  | Ref. |  |  |  |  |
| Not in union | 55.5 | 51.7 | 59.2 | 2.26 | 1.61 | 3.18 | **<0.001** | 1.25 | 0.80 | 2.05 | **0.366** |  |
| Parity at index birth |  |  |  |  |  |  |  |  |  |  |  |  |
| No previous birth | 66.2 | 62.2 | 69.9 | 1.81 | 1.52 | 2.17 | **<0.001** | 1.43 | 1.11 | 1.84 | **0.006** | **0.019** |
| 1 to 2 previous births | 51.9 | 47.6 | 56.2 | Ref. |  |  |  | Ref. |  |  |  |  |
| 3 or more births | 50.8 | 42.3 | 59.4 | 0.96 | 0.62 | 1.35 | **0.807** | 1.33 | 0.89 | 1.99 | **0.163** |  |
| Exposure to media (radio) |  |  |  |  |  |  |  |  |  |  |  |  |
| Not exposed to media | 53.9 | 48.7 | 58.9 | Ref. |  |  |  | Ref. |  |  |  |  |
| Exposed to media | 60.0 | 56.0 | 63.86 | 1.28 | 1.03 | 1.60 | **0.027** | 0.83 | 0.70 | 1.05 | **0.120** |  |
| Antenatal care visits (ANC) |  |  |  |  |  |  |  |  |  |  |  |  |
| No ANC | 12.0 | 7.9 | 17.9 | Ref. |  |  |  | Ref. |  |  |  |  |
| 1-3 ANC | 56.4 | 52.2 | 60.5 | 9.48 | 5.87 | 15.30 | **<0.001** | 8.93 | 5.10 | 15.55 | **<0.001** | **<0.001** |
| 4 plus ANC | 73.8 | 69.8 | 77.4 | 20.63 | 12.6 | 33.90 | **<0.001** | 15.1 | 8.50 | 26.84 | **<0.001** |  |
| Wantedness of index pregnancy |  |  |  |  |  |  |  |  |  |  |  |  |
| Unwanted or mistimed | 57.5 | 51.6 | 63.2 | Ref. |  |  |  | Ref. |  |  |  |  |
| Wanted | 57.6 | 53.7 | 61.4 | 1.01 | 0.77 | 1.31 | **0.966** | 1.41 | 1.00 | 2.07 | **0.084** |  |

Tab3: (continue)
|  | Percentage of facility-based delivery |  |  | Bivariate analysis |  |  |  | Multivariable analysis |  |  |  |  |
| --- | --- | --- | --- | --- | --- | --- | --- | --- | --- | --- | --- | --- |
| Characteristics | % | 95%CI |  | OR | 95%CI |  | Wald test | aOR | 95%CI |  | Wald test | LR test *pvalue |
| Perception of distance to facility |  |  |  |  |  |  |  |  |  |  |  |  |
| Big problem | 43.1 | 38.3 | 48.0 | Ref. |  |  |  | Ref. |  |  |  |  |
| Not a big problem | 69.9 | 66.1 | 73.5 | 3.08 | 2.41 | 3.92 | <0.001 | 1.27 | 1.00 | 1.64 | 0.075 |  |
| Religion |  |  |  |  |  |  |  |  |  |  |  |  |
| Muslim | 53.7 | 50.0 | 57.4 | Ref. |  |  |  | Ref. |  |  |  |  |
| Christian | 83.9 | 77.0 | 89.0 | 4.49 | 2.8 | 7.19 | <0.001 | 2.30 | 1.40 | 3.92 | 0.002 | 0.035 |
| Other | 77.2 | 59.9 | 88.8 | 2.92 | 1.24 | 6.89 | 0.015 | 1.94 | 0.80 | 4.64 | 0.136 |  |
| Region |  |  |  |  |  |  |  |  |  |  |  |  |
| Boké | 45.8 | 37.0 | 54.8 | 1.57 | 0.92 | 2.69 | 0.097 | 1.22 | 0.70 | 2.13 | 0.479 |  |
| Conakry | 86.0 | 79.0 | 90.0 | 11.42 | 6.10 | 21.4 | <0.001 | 1.35 | 0.60 | 2.89 | 0.440 |  |
| Faranah | 38.7 | 30.8 | 47.2 | 1.18 | 0.70 | 1.99 | 0.542 | 1.05 | 0.60 | 1.80 | 0.853 | <0.001 |
| Kankan | 54.4 | 44.1 | 64.3 | 2.22 | 1.26 | 3.93 | 0.006 | 1.58 | 0.90 | 2.65 | 0.084 |  |
| Kindia | 56.3 | 47.0 | 65.1 | 2.40 | 1.40 | 4.13 | 0.002 | 1.36 | 0.80 | 2.31 | 0.248 |  |
| Labé | 34.9 | 26.6 | 44.3 | Ref. |  |  |  | Ref. |  |  |  |  |
| Mamou | 51.8 | 43.1 | 60.4 | 2.00 | 1.18 | 3.40 | <0.001 | 1.64 | 1.00 | 2.74 | 0.058 |  |
| NZérékoré | 77.7 | 68.5 | 84.9 | 6.51 | 3.51 | 12.10 | <0.001 | 4.81 | 2.60 | 8.98 | <0.001 |  |
| Type of place of residence |  |  |  |  |  |  |  |  |  |  |  |  |
| Rural | 85.5 | 81.8 | 88.5 | Ref. |  |  |  | Ref. |  |  |  |  |
| Urban | 44.4 | 39.9 | 48.9 | 7.37 | 5.33 | 10.20 | <0.001 | 2.13 | 1.40 | 3.37 | 0.001 |  |
| Household wealth quintile |  |  |  |  |  |  |  |  |  |  |  |  |
| Poorest | 25.3 | 20.7 | 30.6 | Ref. |  |  |  | Ref. |  |  |  |  |
| Poorer | 46.7 | 40.3 | 53.3 | 2.58 | 1.84 | 3.63 | <0.001 | 1.69 | 1.20 | 2.36 | 0.002 |  |
| Middle | 55.0 | 48.8 | 61.1 | 3.60 | 2.55 | 5.08 | <0.001 | 2.45 | 1.70 | 3.50 | <0.001 | <0.001 |
| Richer | 80.3 | 75.6 | 84.4 | 12.03 | 8.33 | 17.40 | <0.001 | 5.62 | 3.70 | 8.64 | <0.001 |  |
| Richest | 86.7 | 81.5 | 90.7 | 19.27 | 12.00 | 31.00 | <0.001 | 5.83 | 3.20 | 10.70 | <0.001 |  |

\* LR test: Likelihood ratio test

In bivariate analysis, education level, marital status, parity at survey, exposure to media, number of ANC visits, perceived distance to health facility, religion, region, residence, and household wealth quintile were associated with facility-based childbirth in the combined sample of adolescents and young women.

In multivariable analysis (Table 3), those with a secondary or higher educational level were 81% more likely to have given birth in a health facility (95%CI:1.20-2.64) compared to women with no formal education. Compared to those who had no ANC, women who had 1-3 ANC visits (aOR=8.93; 95%CI: 5.10-15.55) and 4 or more visits (aOR=15.1; 95%CI: 8.50-26.84) had higher odds of facility-based childbirth. Christian women had over two-fold higher odds (aOR=2.30; 95%CI: 1.40-3.92) of facility birth compared to Muslim women. Adolescents and young women living in the region of NZérékoré were five time more likely to deliver in a health institution (OR=4.81; 95%CI: 2.60-8.98) compared to women in Labé. Living in an urban area doubled the odds of giving birth in a health facility (aOR=2.13; 95%CI: 1.40-3.37). The likelihood of facility birth increased with each wealthier household quintile. Compared to adolescents and young women living in the poorest household, those in the richest households were nearly 6 times more likely to give birth in a health facility (aOR=5.83; 95%CI: 3.20-10.70). Women who reported having their first birth had 43% higher adjusted odds of facility childbirth compared to those with one or two previous births (p=0.006).

## DISCUSSION

This study examined recent levels and determinants of facility-based childbirth among adolescents and young women in the Republic of Guinea. The overall facility-based childbirth level found was 58%, comparable among adolescents and young women. The study also found that about 6% of women included in the analysis gave birth in the private sector. This was more frequent in young women than adolescents. In addition, most adolescents and young women had initiated prenatal visits, although few reached the recommended four or more ANC visits (37% of the combined sample). In multivariable analyses, education level, number of ANC visits, religion, residence, administrative region, and household wealth index were associated with facility-based delivery. The strongest predictors of higher facility-based childbirth were the use of 4 or more ANC visits, household wealth, and residence in the NZérékoré region. In addition, compared to respondents who were having their second or third child, women giving birth to their first child had higher adjusted odds of giving birth in a health facility.

The overall levels of facility-based childbirth among adolescents and young women found in this study (58%) were statistically higher than what was reported from the same DHS among all women of reproductive age in Guinea (53%). In Guinea, one of the most important policies improving access to maternal health services during this period is the 2011 user fee removal policy [21]. As a result, the percentage of all births occurring in health facilities has increased from 40% in 2012 to 53% in 2018. This increase is even more pronounced in the under 20 years age group (from 41% to 58%, p<0.001) [25,26]. A similar increase has previously been noted in other sub-Saharan African countries, including Uganda [22]. However, despite this increase, the prevalence of facility-based childbirth among adolescents and young women in Guinea is far behind the average for sub-Saharan African countries (65%) [9], including neighboring countries such as Senegal (8%) and Mali (71%) [23,24]. Furthermore, we believe that this result is sub-optimal in light of the challenges associated with high maternal mortality and morbidity in the country and is hampering the achievement of Sustainable Development Goal 3 by 2030.

Our study identified large inequalities in the use of health facilities for childbirth across household wealth quintiles and geographic regions. Compared to adolescents and young women living in the poorest households, those from richer and richest wealth quintiles were more than five times as likely to deliver in a health facility, adjusted for other determinants. This suggests that despite the user fee removal policy, women and their families continue to face substantial financial barriers to accessing childbirth care, potentially including payment of informal fees and the indirect costs of seeking care, such as transportation, food, and sometimes housing. These findings are consistent with reports from other sub-Saharan African countries [7,25,26]. In regard to geographical inequalities, we found that while adolescents and young women living in Conakry had the highest bivariate odds of facility-based delivery, it was the region of NZérékoré in which the highest adjusted odds were found (compared to Labé). While an in-depth understanding of the reasons for this finding, we suspect that local health system actors are implementing national guidelines differently across the country, depending on local challenges such as the availability of financial and human resources. Similar disparities have been reported in Burkina Faso, Niger, Nigeria, Ghana, and Senegal [27].

Next, we draw attention to the fact that the increasing number of births occurring in health facilities, among all women in general, and adolescents and young women in particular, might have consequences for the quality of care provided at childbirth without a concomitant increase in resources. Current challenges of the health sector include health infrastructure, staffing, and funding. For instance, in 2019, only 51% of the public sector health facilities had an adequate physical state of functionality [17]. According to a Guinean Ministry of Health report from 2014, there were approximately 108 obstetrician-gynecologists, 409 midwives, and 1189 nurses for the population of almost 12 million [28]. While our study did not assess the quality of care women received during childbirth, we report patterns consistent with concerns about the quality of care. For example, while only a small percentage of women 15-24 used private sector facilities for childbirth (6%), this level was twice as high among young women compared to adolescents, which might be partly related to previous experience with maternal health services (young women have on average higher parity). Previous research in Guinea showed that lack of staffing, equipment, and space within health facilities generates a negative feedback loop resulting in high levels of disrespect and abuse during childbirth in health facilities. This included women being slapped by attendants, being verbally abused for not complying with health workers’ requests, and giving birth on the floor [29]. A study of three urban hospitals reported that nine out of ten episiotomies were performed without consent, and fewer than 10% of women had access to a companion of choice during labour and childbirth [30]. Importantly, this study of four countries, including Guinea, found that young and less educated women had the highest rates of disrespectful care.

Our study findings also have important implications about the continuity of maternal care during pregnancy and childbirth and beyond during the women’s life-course. Most adolescents and young women in the sample reported receiving some ANC, but only a third had the recommended four or more ANC visits (37%). Receiving four or more ANC visits was associated with 15 times higher adjusted odds of childbirth in a health facility compared to no ANC. While this association might be partly confounded by higher use of both services by women with increased risks (e.g., twin pregnancies), the findings highlight the fact that timely initiation and contact with ANC during pregnancy is a critical factor for the continuity of maternal care during childbirth and the postnatal period. This is the case for high-quality ANC services as shown in a continuum of care analysis from Guinea [31]. ANC is an opportunity for a woman to discuss what her pregnancy implies for herself and her child with a skilled provider, ask questions, and develop a relationship of trust with the health system. It is also an opportunity for the providers to detect potential risk factors and educate the woman on the benefits of continued health service utilization[32].

We found that primiparity was associated with a higher facility-based delivery. This is consistent with studies conducted in sub-Saharan African contexts [33]. One reason for this could be the perceived higher health risk attributed to the first pregnancy in such contexts [34]. We hypothesize that after the first delivery in a facility where the woman may experience poor quality services and mistreatment, she would choose to deliver in a private facility, if she affords, or within her community for the subsequent deliveries [35]. However, a study on facility-based delivery including DHS from 34 sub-Saharan African countries reported that among women having their first birth, the youngest had significantly lower likelihood of delivering in a facility [36]. In any case, experiencing poor quality disrespectful care at the beginning of reproductive life carries negative consequences for the woman’s future use of health services, her wellbeing and survival, and that of her child. This is especially the case in Guinea where the total fertility rate of 4.8 [37] and high maternal mortality rate (576/100,000 live births in 2017) combine into a very high lifetime risk of maternal death of 1 in 35 [1].

### Limitations

This study benefited from a recent nationally representative sample of women of childbearing age. However, we note some limitations. First, the study’s cross-sectional nature does not allow for a temporal relationship between the independent variables and the outcome of interest. Second, there is also the possibility of recall bias that characterizes the DHS, resulting from the retrospective nature of the information collected. Third, because some variables such as ANC were only available for the most recent live birth, we analyzed this subset of births. This might have resulted in underrepresenting the experience and determinants of women with higher number of births in the recall period.

## CONCLUSIONS

This study showed facility-based births among adolescents and young women in Guinea are higher than in the general population. In the pathway to achieve the SDG targets by 2030, there is a need to address inequalities related to financial and geographic accessibility given that the poorest women are still unable to afford the costs of childbirth in health facilities. Also, positive lessons for the regions achieving higher levels should be documented and scaled up. Another gap identified in this study relates to the quality of care across the continuum of care, which requires more investment and accountability in the health sector. Extending beyond the care received at childbirth, attention to accessible, acceptable, affordable, and high-quality maternal care must be paid to antenatal care as the first step in the continuum of care and an important determinant of the use of facilities for childbirth. This is particularly important among adolescents and young women whose well-being will be affected by the care experience and health outcomes experiences at the beginning of their reproductive lives.

## Data Availability

The datasets analyzed during the current study are accessible on https://dhsprogram.com/data/available-datasets.cfm

## Author Contributions

Conceptualization: Fassou Mathias GROVOGUI, Hawa MANET, Alexandre DELAMOU, LenkaBENOVA,

Data curation: Fassou Mathias GROVOGUI, Lenka BENOVA

Formal analysis: Fassou Mathias GROVOGUI, Lenka BENOVA

Writing – original draft: Fassou Mathias GROVOGUI

Writing – review & editing: Fassou Mathias GROVOGUI, Lenka BENOVA, Sidikiba SIDIBE, Nafissatou DIOUBATE, Bienvenu Salim CAMARA, Abdoul Habib BEAVOGUI, Alexandre DELAMOU.

## Notes

### Competing Interest Statement

The authors have declared no competing interest.

### Funding Statement

The author(s) received no specific funding for this work.

